# Estimation of the Final Size of the COVID-19 Epidemic in Pakistan

**DOI:** 10.1101/2020.04.01.20050369

**Authors:** F. Syed, S. Sibgatullah

## Abstract

The COVID-19 infections in Pakistan are spreading at an exponential rate and a point may soon be reached where rigorous prevention measures would need to be adopted. Mathematical models can help define the scale of an epidemic and the rate at which an infection can spread in a community. We used an SIR model to predict the magnitude of the COVID-19 epidemic in Pakistan and compared the numbers with the reported cases on the national database. Our results indicate that Pakistan could hit peak number of infectious cases on May 26th, 2020 and by June 24th, 2020, 90% of the population will have become infected with the virus if policy interventions seeking to curb this infection are not adopted aggressively.

## 1. Introduction

The first case of COVID-19 emerged in Pakistan on 26^th^ February 2020 in Karachi, the most populous city of Pakistan. The patient zero had a travel history to Iran and immediately quarantined upon testing positive for the virus. However, the patient was followed by hundreds of pilgrims returning from Iran which were likely carrying the virus that ultimately led to the spread of COVID-19 into the community. Since then the infections have been increasing exponentially and without proper intervention the situation may escalate enough to overwhelm the already struggling healthcare system in the country.

Since the emergence of COVID-19, several mathematical models have been developed to simulate the rate of infection spread, infections per day and the resolution of the epidemic. The SIR model refers to the number of susceptible, infected and recovered cases during an epidemic at any given time. The model assumes that susceptible cases (S), infected cases (I) and recovered cases (R) are compartments and each individual of a given population will pass through the susceptible phase then to the infected phase and finally to the recovered phase. The SIR model is a steady state model, therefore the population that is analysed is static i.e. no one is being born or is dying. Additionally, the model assumes that once a person is infected, they are immune to the disease and therefore cannot contract it again. The SIR model is ideal for modelling the spread of diseases spread through person to person contact.

In this study we aim to model the COVID-19 epidemic in Pakistan, which will indicate the peak infection day, rate of increase of infections per day and a 90-day forecast to assess the final size of the epidemic.

## 2. Model

The model is built on the following set of assumptions, based on the methodology described by Ronald Ross and William Hammer ^1^, is expressed as the following differential equations:

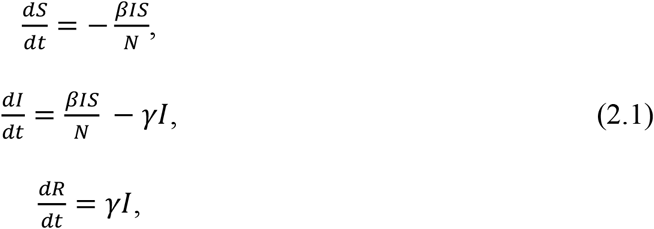

The basic assumption of the SIR model is that the total number of susceptible infected and recovered cases at any given time is equivalent to the test population, so the equations can be represented as:

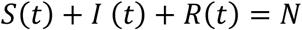

The basic reproduction number (R_°_), is a ratio between the fraction of individuals susceptible per day (β) and the fraction of recoveries (γ); represented as:

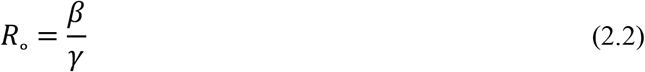

The value of R_°_ plays a significant role in determining the infectiousness of a certain disease-causing organism. Therefore, the rate of change in infected individuals is directly dependent on the R_°_, given by:

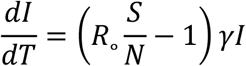

Further assumptions of the SIR model assume that if the R_°_ is greater than the ratio of total population and the susceptible cases at time zero then it would imply that the outbreak will turn into a full-fledged epidemic.

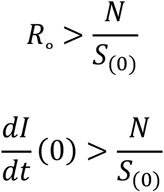

Similarly, if the R_°_ is less than N/S (0), then it would imply that the outbreak will not cause an epidemic. Therefore, the R_°_ plays a crucial role in determining the fate of an epidemic.

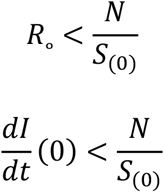

## 3. Data

The data was extracted from daily situation reports published on the National Institute of Health (NIH) Pakistan for a period of 33 days, (from 26 February 2020 to 28^th^ March 2020) and was corroborated with the simulation results.

## 4. Model Parameters

In the case of COVID-19, the value of R_°_ is highly variable and varies from country to country. Several sources ^2–7^ report a range of R_°_ values, between 1.4 – 3.9, therefore we took an average value of 2.65 for our current analysis. The value of R_°_ will continue to evolve as the epidemic progresses throughout the globe; till then, we do not have an exact figure ^2^. The value of γ was considered based on the average infectious period for COVID-19, so γ=0.14. The value of β was calculated to be 0.378 from equation 2.2.

The S_0_ was assumed to be 220,000,000 since the entire population of Pakistan is susceptible to COVID-19, as the disease is new and is spreading across all regions of the country.

The initial time T was set to be day 1, when the first 2 cases of COVID-19 arrived in Karachi. The simulation was used to predict the number of susceptible, infected and recovered cases for a period of 120 days starting from the day the first cases arrived.

## 5. Results

The NIH Pakistan data for the cumulative cases and daily reported cases was plotted to observe the increase in number of cases for a period of 33 days. The current trajectory suggests an exponential increase in the number of cases since the epidemic started in Pakistan on 26^th^ February 2020 (figure 5.1). The data on the number of daily reported cases also reflects a general increasing trend as testing for COVID-19 gathers pace (figure 5.2).

**Figure 5. 1.**
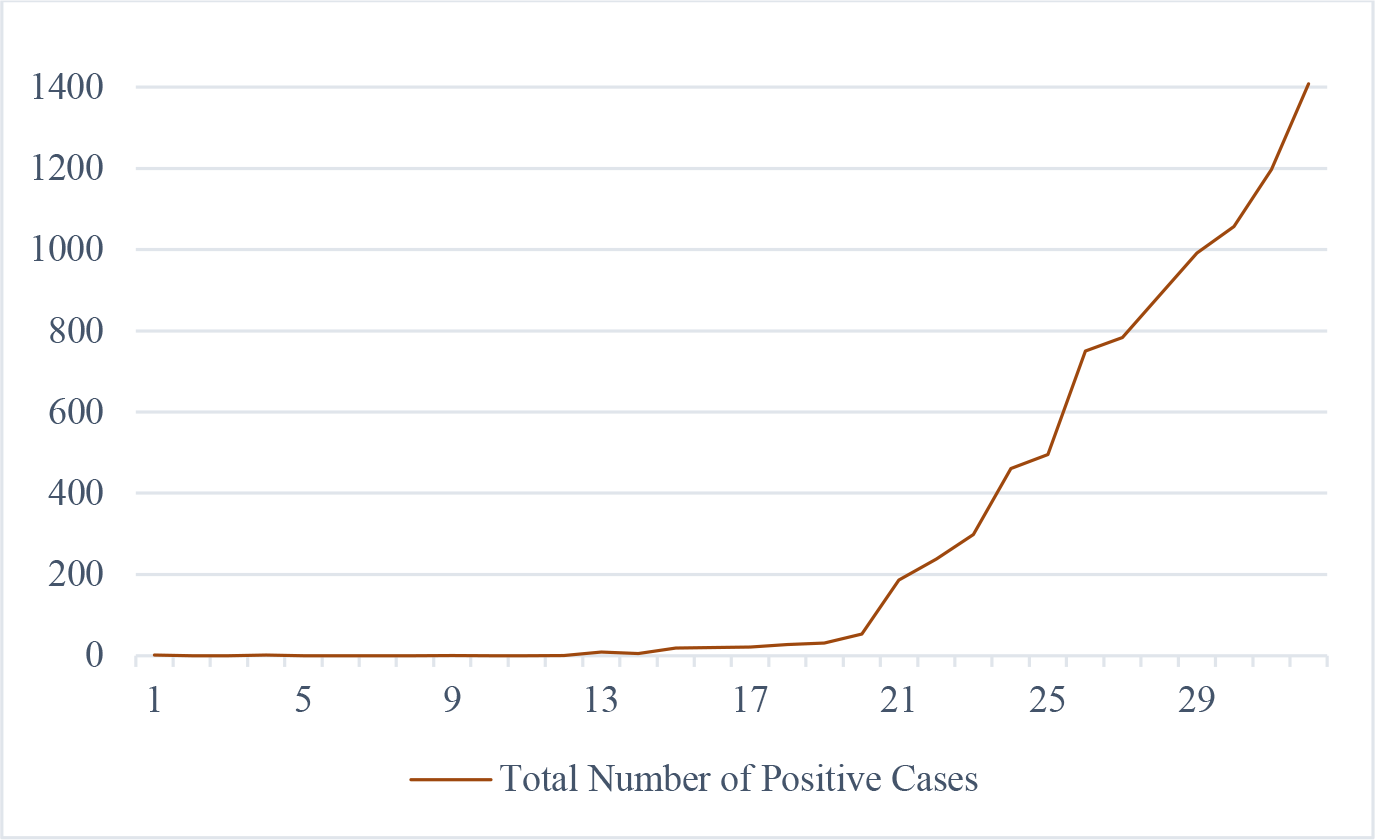
The cumulative COVID-19 cases in Pakistan since the outbreak began on 26^th^ February 2020. The graph shows an exponential growth pattern for the 32-day data extracted from the NIH, Pakistan database. The outbreak started from 2 cases and on day 32 of the epidemic, the number of cases stand at 1408. The x-axis corresponds to the days, whereas the y-axis corresponds to the number of cases.

**Figure 5. 2.**
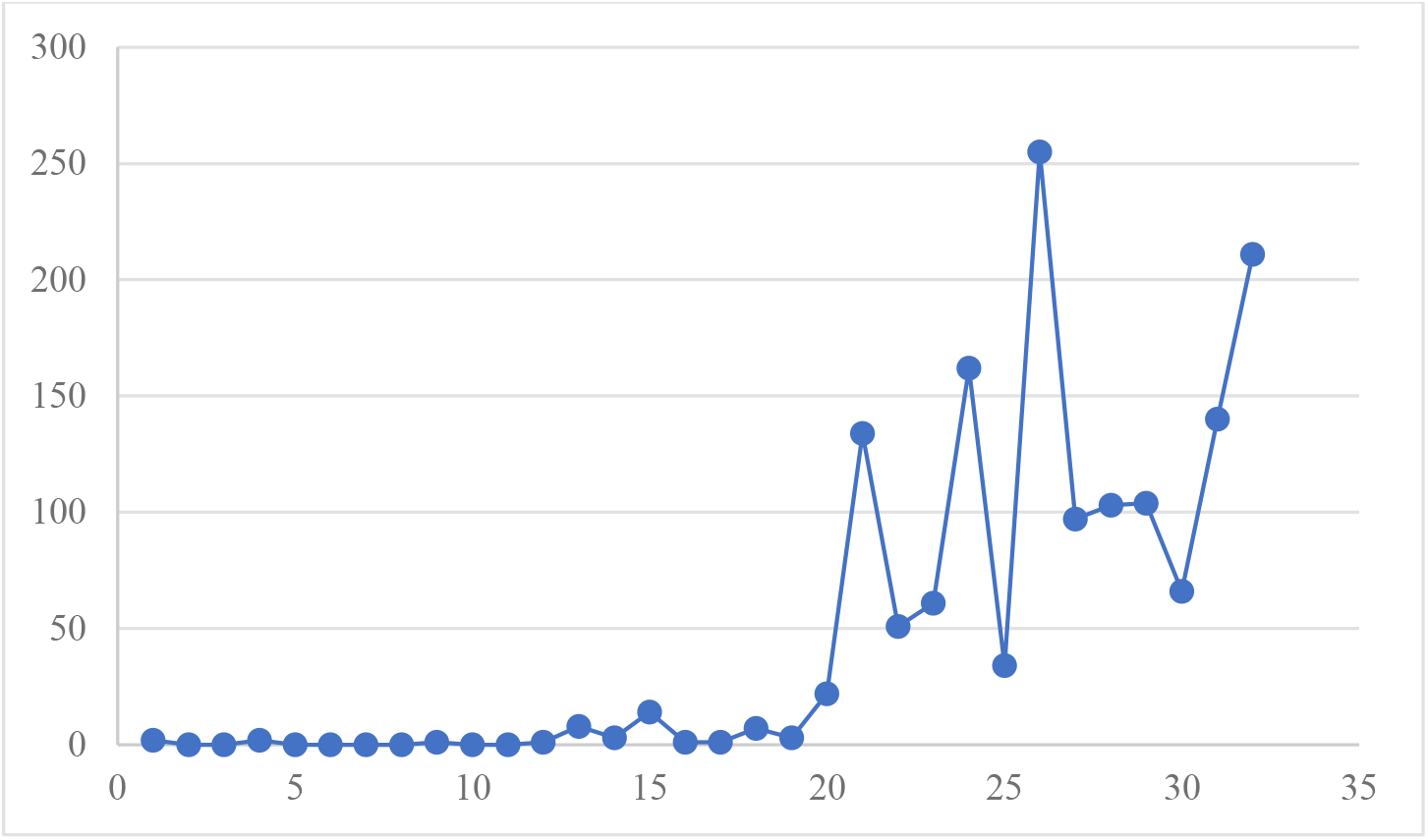
Daily reported COVID-19 cases in Pakistan since the outbreak on 26^th^ February 2020. The graph shows an exponential growth pattern for the 32-day data extracted from the NIH, Pakistan database. On day 1 of the epidemic the number of cases were 2 and on day 32 the number of daily reported cases 211. These numbers may represent the gap in the unreported or asymptomatic cases in Pakistan. The x-axis corresponds to the days, whereas the y-axis corresponds to the number of cases.

The SIR model for the spread of COVID-19 in Pakistan (figure 5.3), under the assumptions mentioned in Model Parameters, indicate that the number of infections will peak on Day 91 (26^th^ May 2020), where 5,921,1209 individuals could be potentially infected. The number of infections will rapidly decrease and on the 120^th^ day (24^th^ June 2020) the number of infected cases will stand at 369, 8192, whereas 198,761,399 cases will have recovered which is approximately 90% of the Pakistani population. We also superimposed the modelled data with the reported numbers (figure 5.4). Our calculations suggest that a significant correlation (r=0.97, p<0.001) between the modelled trajectory the actual number of reported cases. Therefore, the current values for R_°_, β and γ seem to be accurate for the population of Pakistan.

**Figure 5. 3.**
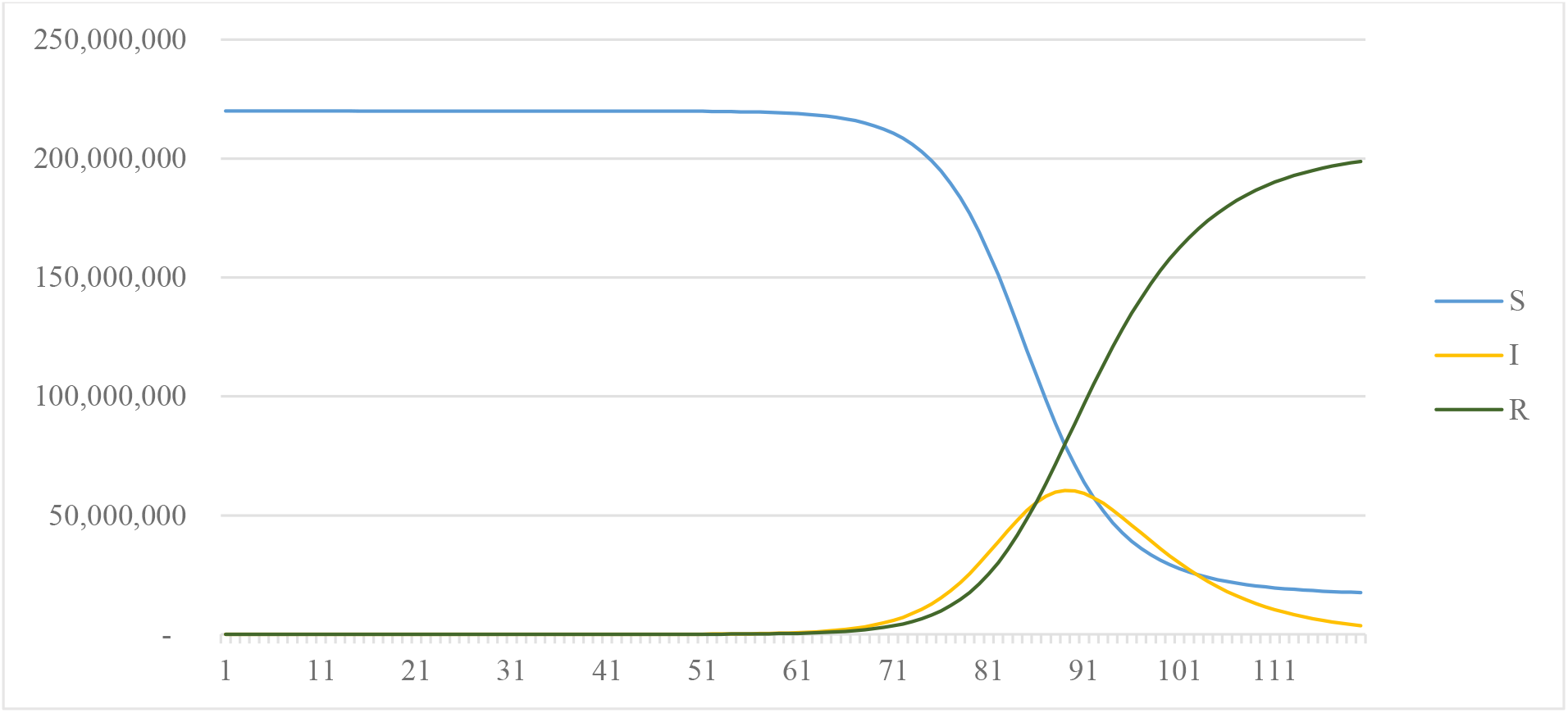
The SIR Model of the COVID-19 Epidemic in Pakistan. The simulations suggest that peak infection day will fall on T=91, where I_(91)_ =5, 921, 1209. The epidemic should have resolved by T=120 where the value of R_(120)_ =198,761,399.03. Susceptible cases (S) shown in Blue, Infected cases (I) shown in Yellow and Recovered cases (R) shown in Green. The x-axis represents the number of days, whereas the y-axis represents the number of cases.

**Figure 5. 4.**
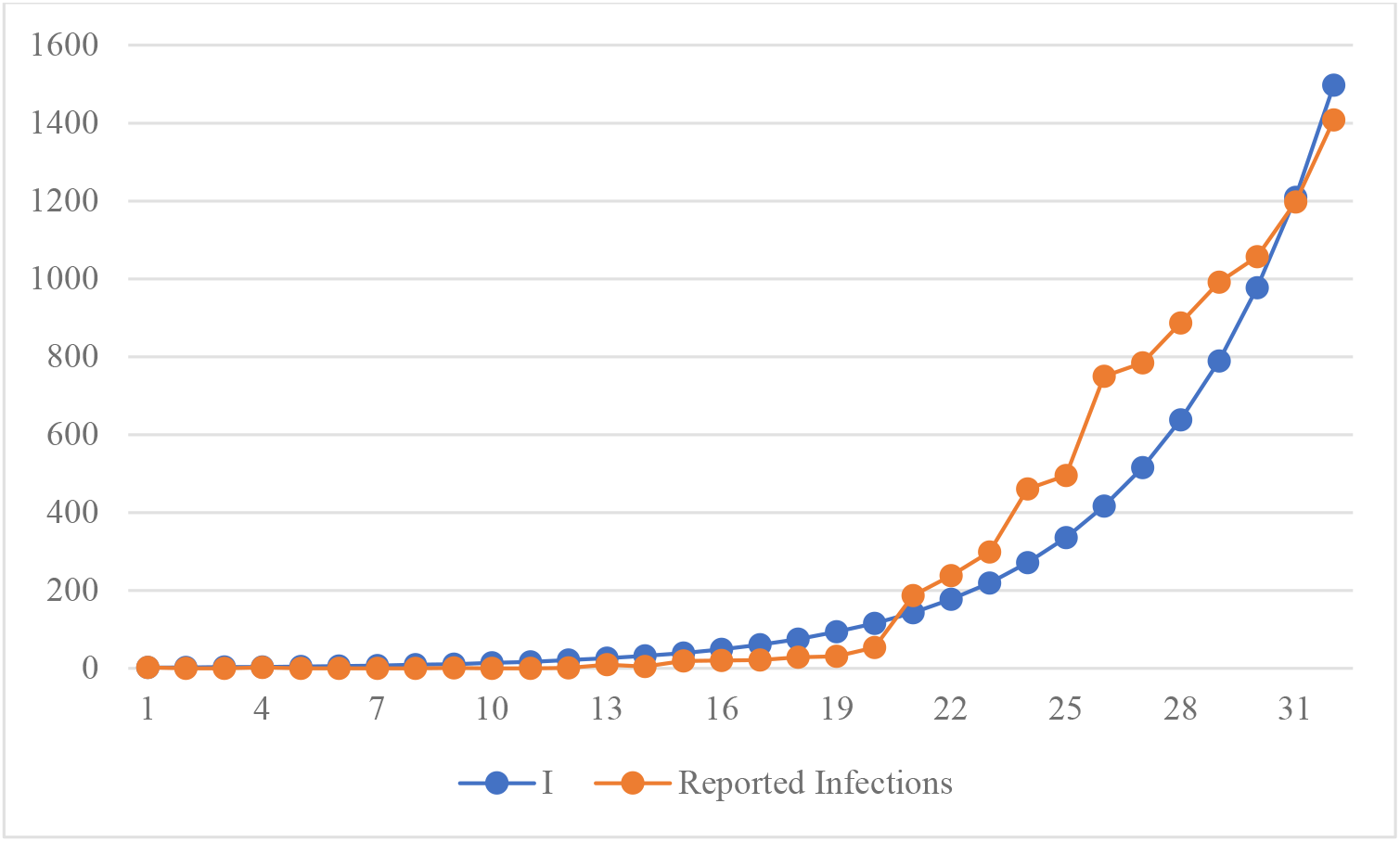
Comparison of Reported data with the Modelled data. The modelled data for infected cases (I) seems to follow the trajectory of the reported data from the NIH Pakistan database. We found a strong correlation between the trends of the modelled data (I) and the reported data (r=0.97, p<0.001). The modelled data is represented as a Blue line and the reported data is represented as an Orange line. The x-axis represents the number of days, whereas the y-axis represents the number of cases.

## 6. Discussion

Our study was focused on modelling the COVID-19 epidemic in Pakistan in order to estimate the number of infections, the peak infection day, the rate of increase of infections per day and the resolution of the end-point of the epidemic ^1^.

The simulation parameters were adjusted according to the population of Pakistan. Our model simulated the conditions where COVID-19 is spreading in a closed population of 220,000,000 people, without the effect of any extraneous variables such as social distancing, hand washing or travel restrictions. The values of R_°_=2.65, β=0.378 and γ=0.14 generated data which was quite close to the actual reported cases in Pakistan (figure 5.4). Furthermore, the trends of the modelled I and the reported number of infections coincide significantly (r=0.97, p<0.001). Thus, we can safely assume that these results can be used as a predictor of a likely scenario of COVID-19 for Pakistan.

According to the simulations the peak infection day will occur on 26^th^ May 2020, where 5,921,1209 persons could get infected with the virus in a single day. Previous reports from China ^8^ have indicated that COVID-19 initially follows an exponential growth pattern ^9^ coupled with asymptomatic carriers ^10^, leads to a rapid increase in the number of infections. Correlating the reported data with our simulations, we can assume that if left unchecked, the disease can spread at an unprecedented rate in Pakistan, especially in closely knit communities and densely populated areas.

The results of our study also predict that within a time span of 120 days, 90% of the population will have been infected with COVID-19. These include asymptomatic carriers, which could account for approximately 20% of the population ^11^ and will have the tendency to unknowingly infect others. However, the major concern for Pakistan would be the healthcare system which would not be able to cope with the overwhelming number of patients if the trajectory remains the same. Studies place the mortality rate of COVID-19 at 2.3%, severe cases at 14% and critical cases at 5% ^12,13^, which would imply that potentially 50,000 could die; 30,800,000 cases could become severe and 11,000,000 could become critical during the aftermath of epidemic in Pakistan. Therefore, there is an urgent need to implement effective measures to curb the rise in COVID-19 infections in Pakistan, otherwise it could lead to drastic consequences. These measures could decrease the value of β (the fractions of individuals susceptible per day) and in turn would decrease the R?(the basic reproduction rate) of the virus, (see equation 2.2).

Our study gives a well-balanced estimate of how the COVD-19 could spread in Pakistan. However, the study has a few limitations, such as the effect of extraneous variables has not been considered in the modelling. This could also serve as a future direction for us to model an epidemic where these factors come into play.

In conclusion, the SIR modelling of the COVID-19 in Pakistan revealed that the infection could spread at an exponential rate if proper measures are not taken to reduce its transmission, through safe practices such social distancing, hand-washing and large scale testing of suspected cases in the region.

## Data Availability

Kindly contact the corresponding author for additional data.

## References

1. Hethcote HW. Mathematics of infectious diseases. SIAM Review. 2000;42(4):599–653. doi:10.1137/S0036144500371907

2. Wu JT, Leung K, Bushman M, et al. Estimating clinical severity of COVID-19 from the transmission dynamics in Wuhan, China. Nature Medicine. March 2020:1–5. doi:10.1038/s41591-020-0822-7

3. Li Q, Guan X, Wu P, et al. Early Transmission Dynamics in Wuhan, China, of Novel Coronavirus–Infected Pneumonia. New England Journal of Medicine. 2020;382(13). doi:10.1056/nejmoa2001316

4. Riou J, Althaus CL. Pattern of early human-to-human transmission of Wuhan 2019 novel coronavirus (2019-nCoV), December 2019 to January 2020. Euro surveillance: bulletin Europeen sur les maladies transmissibles = European communicable disease bulletin. 2020;25(4). doi:10.2807/1560-7917.ES.2020.25.4.2000058

5. Liu T, Hu J, Kang M, et al. Time-varying transmission dynamics of Novel Coronavirus Pneumonia in China. bioRxiv. February 2020:2020.01.25.919787. doi:10.1101/2020.01.25.919787

6. Wang H, Wang Z, Dong Y, et al. Phase-adjusted estimation of the number of Coronavirus Disease 2019 cases in Wuhan, China. Cell Discovery. 2020;6(1):1–8. doi:10.1038/s41421-020-0148-0

7. Read JM, Bridgen. Properties of AdeABC and AdeIJK efflux systems of Acinetobacter baumannii compared with those of the AcrAB-TolC system of Escherichia coli. Intergovernmental Panel on Climate Change, ed. Antimicrobial agents and chemotherapy. 2014;58(12):7250–7257. doi:10.1101/2020.01.23.20018549

8. Wu Z, Jama JM-, 2020 undefined. Characteristics of and important lessons from the coronavirus disease 2019 (COVID-19) outbreak in China: summary of a report of 72 314 cases from the Chinese. jamanetwork.com. https://jamanetwork.com/journals/jama/article-abstract/2762130.

9. Liu Y, Gayle A, … AW-S-J of travel, 2020 undefined. The reproductive number of COVID-19 is higher compared to SARS coronavirus. academic.oup.com.

10. Bai Y, Yao L, Wei T, et al. Presumed asymptomatic carrier transmission of COVID-19. jamanetwork.com. https://jamanetwork.com/journals/jama/article-abstract/2762028.

11. Mizumoto K, Kagaya K, Zarebski A, Chowell G. Estimating the asymptomatic proportion of coronavirus disease 2019 (COVID-19) cases on board the Diamond Princess cruise ship, Yokohama, Japan, 2020. Eurosurveillance. 2020;25(10). doi:10.2807/1560-7917.es.2020.25.10.2000180

12. Ji Y, Ma Z, Peppelenbosch MP, Pan Q. Potential association between COVID-19 mortality and health-care resource availability. The Lancet Global Health. 2020;8(4):e480. doi:10.1016/S2214-109X(20)30068-1

13. Wu Z, McGoogan JM. Characteristics of and Important Lessons from the Coronavirus Disease 2019 (COVID-19) Outbreak in China: Summary of a Report of 72314 Cases from the Chinese Center for Disease Control and Prevention. JAMA - Journal of the American Medical Association. 2020. doi:10.1001/jama.2020.2648

